# Childhood adversities and accident mortality in early adulthood - a population-based cohort study

**DOI:** 10.64898/2026.02.18.26346533

**Authors:** Line Marie Toft Dyhr, Naja Hulvej Rod, Leonie Elsenburg

## Abstract

Childhood adversities are common and linked to increased risk of premature mortality, including deaths from accidents in early adulthood. We examined associations between childhood adversity and specific types of lethal accidents using nationwide register data from 1,282,636 individuals in the DANish LIFE course (DANLIFE) cohort born between Jan 1, 1980, and Dec 31, 2001, who did not die or emigrate before age 16. Individuals were classified into five trajectory groups based on annual exposure to 12 adversities across three dimensions from ages 0–15. Accident mortality was categorised into traffic, narcotic and hallucinogenic, other poisoning, and other accidents. Individuals were followed through Dec 31, 2022. Relative and absolute risks were estimated using Cox proportional hazards and Aalen additive hazard models.

Compared with the low-adversity group, individuals in one of the childhood adversity groups experienced 4.4 to 33.8 additional accident deaths per 100,000 person-years. The largest relative (HR=13.4 95% CI [9.9–18.6]) and absolute (HD=12.9 95%CI [10.0–15.8]) differences were identified for the high versus low adversity group. High childhood adversity is strongly associated with preventable accident mortality in early adulthood, underscoring the need for structural and social interventions to reduce adversity exposure and related excess mortality.

## Introduction

Exposure to childhood adversities is common. Childhood adversities cover a vast range of childhood factors, events and circumstances, such as family dysfunction, poverty, severe illness in the family, substance abuse and parental divorce^1–4^. Even in a country with extensive social security, such as Denmark, one in ten children have experienced more than three adversities between early infancy and late adolescence^5^. Similar or more extreme prevalences of childhood adversities have been found across countries. A systematic review and meta-analysis found that on average 13% of the total sample, mainly representing estimates from the US and UK, had experienced at least four childhood adversities^1^.

Early life disadvantage such as experiencing childhood adversities, has long been recognized as an important determinant of later life health, morbidity and mortality^3^. Exposure to one childhood adversity also often precipitates additional adversities, leading to a cumulative burden of adversities^6,7^. An extensive body of literature has shown that those who grow up experiencing adversities have a higher risk of premature mortality^1,3,4,8–15^. We previously found that accidents were the most common cause of premature mortality in the Danish population aged 16-35 years, accounting for 37% of deaths. Furthermore, we found that children who had experienced childhood adversities had a 2-4.5 times higher risk of dying from accidental causes of death in early adulthood compared with children who experienced low childhood adversity^4^. Previous studies of childhood adversities, combined or in isolation, on all-cause and cause-specific mortality confirm these findings^1,3,4,8–15^.

However, studies examining the association between childhood adversities and different types of accidental causes of death are lacking. By investigating different accident types, we acknowledge that accidental causes of death are diverse and thus have different points for intervention and prevention. The existing literature mainly focused on the relation between childhood adversities and mortality caused specifically by traffic accidents or accident mortality together with other external causes of mortality such as homicide and suicide^8,9,14–17^. In order to inform future preventive strategies to lower the incidence of accident mortality in early adulthood, it is however paramount to investigate what types of accidents are most common and which subgroups of the population are at higher risk of dying from these specific types of accidents.

Another added value of this study is that we employ a measure of childhood adversities using based on high resolution data from high quality registries, allowing us to account for multiple and recurring childhood adversities as well as aspects of both timing and accumulation. In this study, we will investigate the risk of different lethal accident types across trajectory groups of adversities, as this will provide valuable insight into the nature of the association across accident types. Using unselected nationwide register data with objective and repeated information on childhood adversity exposure and early adulthood accident mortality, we aim to identify the most common lethal accident types and estimate how the risk of mortality from these accident types varies across the different trajectory groups of childhood adversity. The accident types we will examine are traffic accidents, narcotics and hallucinogenics accidents, other poisoning accidents and other accidents.

## Methods

### Study population

The study is based on data the Danish life course (DANLIFE) cohort which contains information from multiple nationwide registries in Denmark^5^. Access to pseudonymized, securely stored data was granted by Statistics Denmark and the Danish Health Data Authority. The use of unique personal identification numbers assigned to all Danish residents enabled individual-level linkage across registers, including linkage between parents and children^18^. The DANLIFE cohort includes 2,223,927 individuals born between 1 January 1980 and 31 December 2015^5^. In this study, we excluded individuals born after 31 December 2001 (N=866,119), those who emigrated before age 16 (N=61,355), those who died before age 16 (N=12,498), and those with missing information on covariates (N=1,319) included in the main analysis. The final study population consists of 1,282,636 individuals (flowchart in Supplementary Figure S1). The DANLIFE study received approval from the Danish Data Protection Agency via the joint notification submitted by the Faculty of Health and Medical Sciences, University of Copenhagen (record no. 514-0641/21-3000). In Denmark, registry studies are exempt from ethical approval.

### Childhood adversities

This study uses the childhood adversity measure developed by Rod et al. (2020). Exposure to childhood adversities was assessed based on allocation to one of the five most common trajectory groups that model exposure to adversities across three dimensions of childhood adversity; material deprivation (i.e. family poverty and parental long-term unemployment), loss or threat of loss (i.e. death of a parent, death of a sibling, parental somatic illness and sibling somatic illness), and family dynamics (i.e. foster care, parental or sibling psychiatric illness, parental alcohol or drug abuse, maternal separation). These dimensions were identified by a panel of experts in child health, stress and child psychology after thorough examination of the literature^5^. Supplementary material online (Table S1) provides an overview of childhood adversities, their definition and in which registers the information was sourced. The trajectory groups were identified using a group-based multi trajectory model, using the annual rate of adversity exposure to allocate each child to the trajectory group they were most likely to belong to^4^. The five trajectories are ‘low adversity’, ‘early material deprivation’, ‘persistent material deprivation’, ‘loss or threat of loss’, and ‘high adversity’. Characteristics of the trajectory groups are presented in Figure 1. See Supplementary figure S2 for more details.

**Figure 1.**
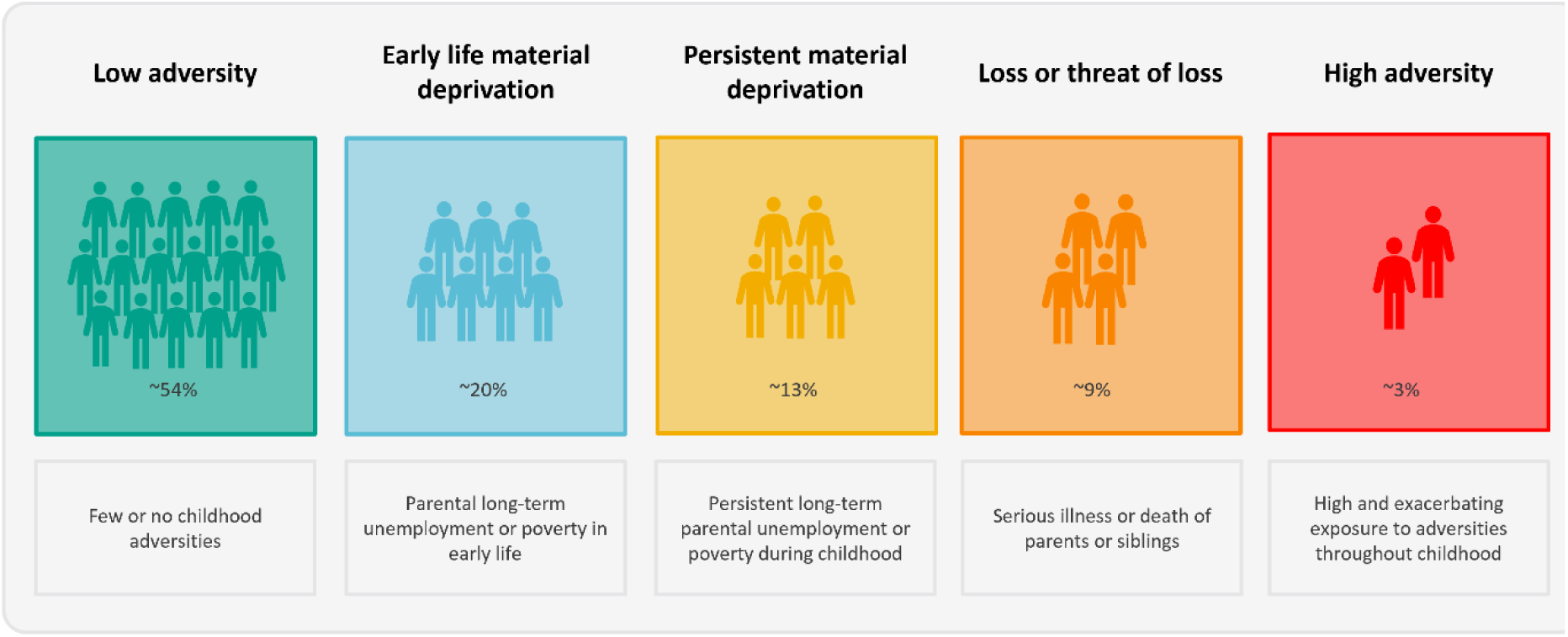
– The five childhood adversity groups and their characteristics. The high adversity group is exposed to high levels of adversity across all three dimensions

### Accident mortality

Information on the accident mortality was obtained through the Cause of Death Register, for which information has been updated until December 31^st^, 2022^19^. Individuals registered after their 16th birthday and before the end of follow-up (31st of December 2022) with an ICD-10 code in the range V01-X59 were considered to have experienced accident mortality. For the remainder of this paper ‘accidents’ refer to these lethal accidents. They were divided into four types: ‘traffic accidents’ (V01-V99), ‘narcotics and hallucinogenics’ (X42), ‘other poisonings’ (X40-X41, X43-X49), and ‘other accidents’ (W00-W99, X00-X39, X50-X59). Other poisonings consisted mainly of accidental deaths from medical drugs, alcohol poisonings and gaseous poisonings (X40-41, X43-X49). Other accidents mainly consisted of drowning, falling, exposure to electricity, or suffocation on gastric content (for details, see supplementary Table S3).

### Covariates

Covariates identified as potential confounders included sex, year of birth, maternal age at birth (<20 years, 20-30 years and >30 years) and parental origin. If one or both parents were from either Europe, North America, Australia or New Zealand, they were registered as of Western origin; otherwise, they were registered as of non-Western origin. Two additional covariates that were adjusted for in sensitivity analyses were parental education and being small for gestational age (SGA). Based on the highest education level attained by either parent at the time of birth, parental education was defined as short (<10 years), medium (10-12 years), and long (12+ years). SGA was defined as having a birthweight below the 10th percentile of the sex-specific standard reference curves for intrauterine growth.

### Statistical analysis

Age-specific incidence rates of the four accident types, separately as well as overall, between age 16 to 42 years were described using Poisson regression models with splines (6 degrees of freedom). Using a Cox proportional hazards model with the childhood adversity trajectory groups as the exposure, we estimated hazard ratios (HRs) and 95% CIs for the four lethal accident types as well as overall accident mortality. Age served as the underlying time scale. Hazard differences (HDs) with 95% CIs were estimated using Aalen’s additive hazard model. The main analysis was adjusted for sex, year of birth, maternal age at birth and parental origin. Individuals were followed from their 16th birthday until death, emigration or end of follow-up on December 31st, 2022, whichever occurred first. In all analyses, the reference group were those who experienced low adversity during childhood. Two sensitivity analyses were made to assess the effects of additional adjustment for parental education and SGA. When numbers allowed, we additionally stratified the analyses by sex.

## Results

Characteristics of the population across trajectory groups are presented in Table 1. In this study, 48.7% of the population were women. The majority of children belonged to the low adversity group, characterized by no or few adversities in childhood (54%), while almost half of the population experienced some level of childhood adversity, with 3% of the population experienced high levels of childhood adversity. In the low adversity group, 8% were born to parents with short education, while this proportion was 54% in the high adversity group. In the high adversity group, 11% were born to young mothers (<20 years), whereas this applied to only 1% of the individuals in the low adversity group. The proportion of children born to parents with non-Western origin was smallest in the low adversity group (1%) and largest in the persistent material deprivation group (8%). During follow-up, 2,366 individuals died from accidents. The most common type of lethal accident was ‘traffic accidents’ (55.9%), followed by ‘narcotics and hallucinogenic accidents’ (15.8%), ‘other poisoning accidents’ (15.7%), and ‘other accidents’ (12.6%). Males were overrepresented in all accident types with the proportion ranging between 80-85% (Supplementary table S2).

**Table 1.** – Background characteristics of the study population at the time of birth across the five childhood adversity trajectory groups.

| **Total** | **Overall**<br>N=1,282,636 | **Low adversity**<br>N=693,870<br>(54.1%) | **Early-life material**<br>**deprivation**<br>N=257,821<br>(20.1%) | **Persistent material**<br>**deprivation**<br>N=171,544<br>(13.4%) | **Loss or threat of loss**<br>N=117,905<br>(9.2%) | **High adversity**<br>N=41,496<br>(3.2%) |
| --- | --- | --- | --- | --- | --- | --- |
| **Sex** |  |  |  |  |  |  |
| Men | 657,972 (51.3%) | 356,005 (51.3%) | 131,842 (51.1%) | 87,629 (51.1%) | 60,029 (50.9%) | 22,467 (54.1%) |
| Women | 624,664 (48.7%) | 337,865 (48.7%) | 125,979 (48.9%) | 83,915 (48.9%) | 57,876 (49.1%) | 19,029 (45.9%) |
| **Maternal age** |  |  |  |  |  |  |
| <20 years | 35,232 (2.7%) | 6,035 (0.9%) | 9,121 (3.5%) | 11,678 (6.8%) | 4,039 (3.4%) | 4,359 (10.5%) |
| 20-30 years | 862,248 (67.2%) | 450,153 (64.9%) | 187,947 (72.9%) | 121,446 (70.8%) | 74,614 (63.3%) | 28,088 (67.7%) |
| >30 years | 385,156 (30.0%) | 237,682 (34.3%) | 60,753 (23.6%) | 38,420 (22.4%) | 39,252 (33.3%) | 9,049 (21.8%) |
| **Parental origin** |  |  |  |  |  |  |
| Non-Western | 35,597 (2.8%) | 7,318 (1.1%) | 9,029 (3.5%) | 14,381 (8.4%) | 4,059 (3.4%) | 810 (2.0%) |
| Western | 1,247,039 (97.2%) | 686,552 (98.9%) | 248,792 (96.5%) | 157,163 (91.6%) | 113,846 (96.6%) | 40,686 (98.0%) |
| **Parental education** |  |  |  |  |  |  |
| Short (<10 years) | 212,958 (16.6%) | 55,391 (8.0%) | 55,066 (21.4%) | 55,453 (32.3%) | 24,758 (21.0%) | 22,290 (53.7%) |
| Medium (10-12 years) | 632,272 (49.3%) | 333,825 (48.1%) | 141,326 (54.8%) | 83,324 (48.6%) | 59,032 (50.1%) | 14,765 (35.6%) |
| Long (>12 years) | 432,741 (33.7%) | 303,084 (43.7%) | 60,455 (23.4%) | 31,518 (18.4%) | 33,667 (28.6%) | 4,017 (9.7%) |
| Missing | 4,665 (0.4%) | 1,570 (0.2%) | 974 (0.4%) | 1,249 (0.7%) | 448 (0.4%) | 424 (1.0%) |
| **SGA** |  |  |  |  |  |  |
| No | 1,071,546 (83.5%) | 592,211 (85.3%) | 214,613 (83.2%) | 137,728 (80.3%) | 96,470 (81.8%) | 30,524 (73.6%) |
| Yes | 168,277 (13.1%) | 79,373 (11.4%) | 35,218 (13.7%) | 26,729 (15.6%) | 17,629 (15.0%) | 9,328 (22.5%) |
| Missing | 42,813 (3.3%) | 22,286 (3.2%) | 7,990 (3.1%) | 7,087 (4.1%) | 3,806 (3.2%) | 1,644 (4.0%) |

The age-specific incident rate of all lethal accidents was highest for both females and males during age 16-19 years, after which the incidence rate decreased until age 24 and then largely plateaued (Figure 2). The high incidence rate in the late teenage years, particularly for males, was mainly driven by traffic accidents (see supplementary Figure S4). The incidence rate patterns for narcotics and hallucinogenics accidents and for other poisoning accidents showed low incidence rates that steadily increased with increasing age. The pattern for other accidents was also similar, but less pronounced and with lower rates. See Supplementary Figure S4 for accident type age-specific incidence rates according to sex.

**Figure 2.**
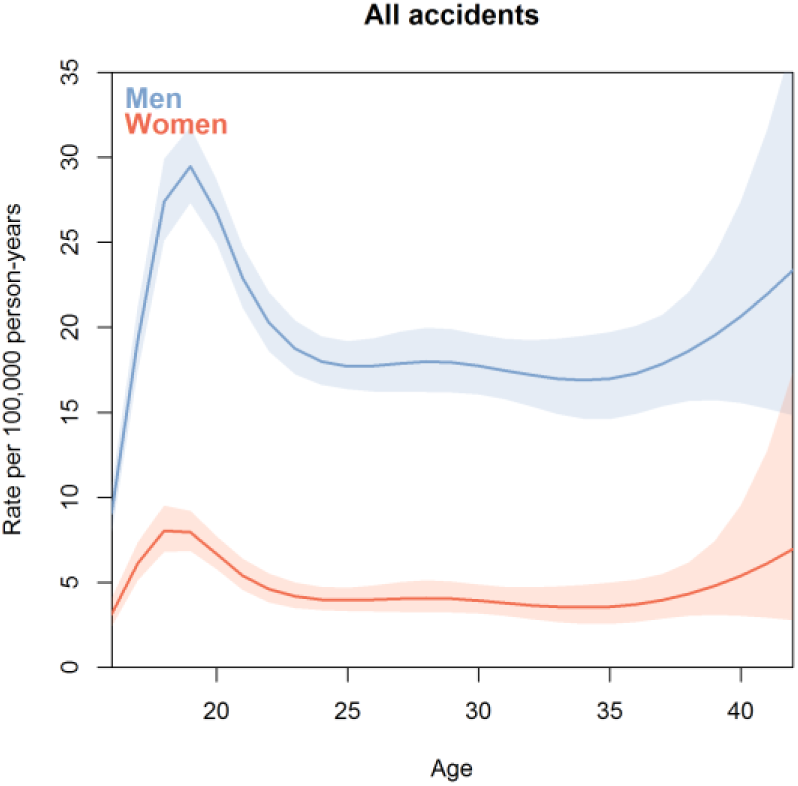
Age-specific incidence rate of accident mortality per 100,000 person-years according to sex.

From the cumulative risk plot, we deemed HRs appropriate to assess the relative risks of lethal accidents across the adversity groups (see Supplementary Figure S3). Compared with the low adversity group, all other adversity groups had a higher number of individuals who died from accidents per 100,000 person-years with HR estimates ranging from HR = 1.52 (95% CI [1.36-1.69]) for the early material deprivation group to HR = 4.55 (95% CI [3.96-5.23]) for the high adversity group. In the high adversity group, this translated to an additional 33.8 deaths (95% CI [28.8-39.0]) per 100,000 person-years compared with the low adversity group (Table 3).

**Table 3.** – Hazard ratios (HR) and hazard differences (HD) per 100 000 person-years with 95% CIs according to childhood adversity trajectory group for overall accident mortality and accident type specific mortality: traffic accidents, narcotics and hallucinogenics accidents, other poisoning accidents and other accidents.

| Accident type | Adversity group | Cases | HR<sup>a</sup> (95%CI) | HD<sup>a</sup> (95%CI) |
| --- | --- | --- | --- | --- |
| All accidents | Low adversity | 817 | 1 (ref) | 0 (ref) |
|  | Early life material deprivation | 526 | 1.52 (1.36-1.69) | 4.4 (3.1-5.7) |
|  | Persistent material deprivation | 490 | 1.80 (1.61-2.02) | 7.2 (5.5-8.8) |
|  | Loss or threat of loss | 255 | 1.89 (1.64-2.17) | 7.5 (5.4-9.6) |
|  | High adversity | 278 | 4.55 (3.96-5.23) | 33.8 (28.8-39.0) |
| Traffic accidents | Low adversity | 512 | 1 (ref) | 0 (ref) |
|  | Early life material deprivation | 326 | 1.51 (1.31-1.73) | 2.8 (1.7-3.8) |
|  | Persistent material deprivation | 277 | 1.63 (1.40-1.89) | 3.6 (2.3-4.8) |
|  | Loss or threat of loss | 123 | 1.45 (1.19-1.76) | 2.4 (0.9-3.8) |
|  | High adversity | 85 | 2.24 (1.78-2.83) | 7.2 (4.3-10.2) |
| Narcotics and hallucinogenics | Low adversity | 88 | 1 (ref) | 0 (ref) |
|  | Early life material deprivation | 64 | 1.70 (1.23-2.34) | 0.6 (0.2-1.1) |
|  | Persistent material deprivation | 85 | 2.92 (2.15-3.96) | 1.8 (1.1-2.5) |
|  | Loss or threat of loss | 48 | 3.31 (2.33-4.70) | 2.1 (1.2-3.0) |
|  | High adversity | 89 | 13.43 (9.94-18.15) | 12.9 (10.0-15.8) |
| Other poisonings | Low adversity | 102 | 1 (ref) | 0 (ref) |
|  | Early life material deprivation | 68 | 1.53 (1.12-2.08) | 0.5 (0.1-1.0) |
|  | Persistent material deprivation | 72 | 2.09 (1.53-2.85) | 1.2 (0.5-1.8) |
|  | Loss or threat of loss | 50 | 2.94 (2.10-4.13) | 2 (1.2-2.9) |
|  | High adversity | 80 | 10.24 (7.59-13.81) | 11.3 (8.5-14.1) |
| Other accidents | Low adversity | 115 | 1 (ref) | 0.0 (ref) |
|  | Early life material deprivation | 68 | 1.40 (1.03-1.89) | 0.5 (0.0-1.0) |
|  | Persistent material deprivation | 56 | 1.47 (1.06-2.05) | 0.6 (0.0-1.2) |
|  | Loss or threat of loss | 35 | 1.82 (1.24-2.66) | 1.0 (0.2-1.7) |
|  | High adversity | 24 | 2.74 (1.75-4.28) | 2.3 (0.8-3.9) |

<sup>a</sup>adjusted for sex, year of birth, maternal age at birth and parental origin.

All adversity groups had a higher risk of traffic accident mortality compared with the low adversity group (Table 3). Looking at lethal accidents from narcotics and hallucinogenics, we observed a more than 13-fold higher risk in mortality in the high adversity group compared with the low adversity group, corresponding to 12.9 (95% CI [10.0-15.8]) additional deaths per 100,000 person-years. Similar estimates across trajectory groups were seen for accident mortality from other poisonings, where a 1.5– to 10-fold higher risk was observed comparing with the low adversity group. The higher risk associated with childhood adversity persisted when looking at other accident types though estimates were lower.

Stratifying the analysis for overall accident mortality on sex, we found no profound difference in relative risk between males and females (Figure 3). However, there were marked differences in the absolute risks between males and females due to the underlying higher risk in men. In the high adversity group, females had 11.6 (95% CI [7.3-16.5]) additional cases of accident mortality per 100,000 person-years, whereas this estimate was 52.7 (95% CI [43.9-61.5]) for males (Figure 3).

**Figure 3.**
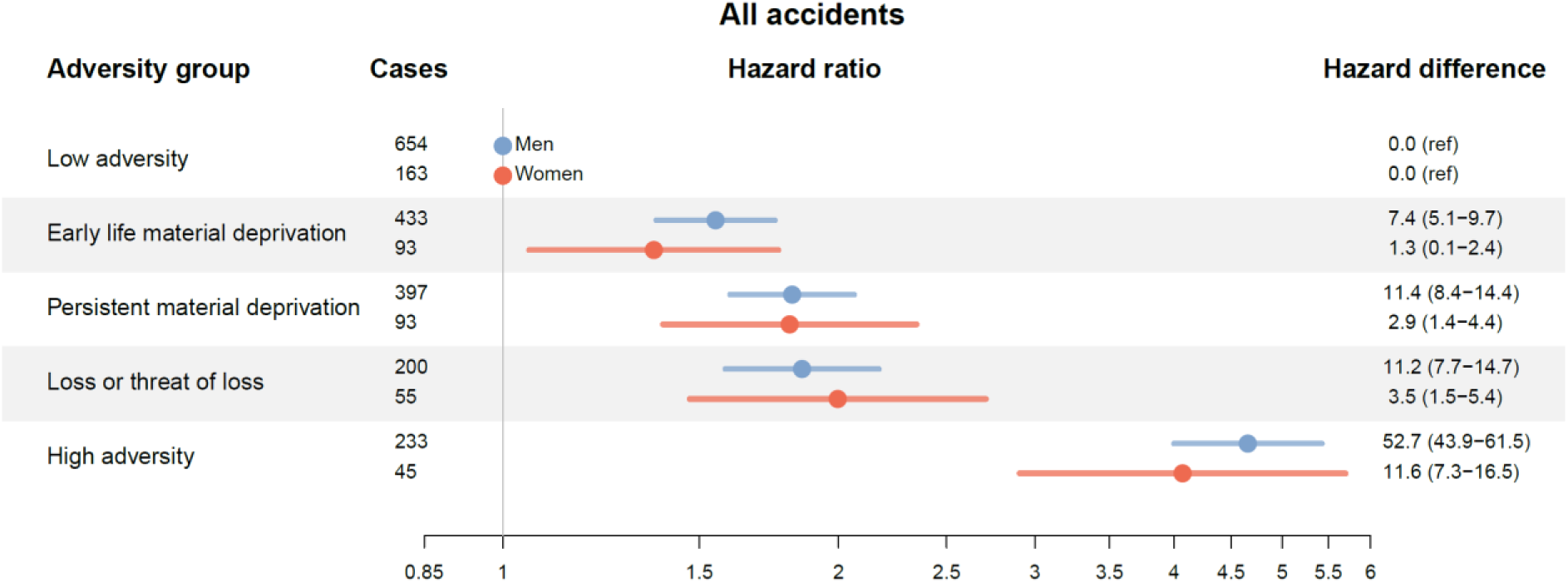
Hazard ratios (HR) and hazard differences (HD) per 100 000 person-years with 95% CIs according to childhood adversity trajectory group for overall accident mortality for men and women.

In the sensitivity analyses we excluded those with missing information on parental education (N=4,665) and those with missing information on SGA (N=42,813), respectively. Additional adjustment for parental education attenuated the risk estimates moderately (Supplementary Figure S5), but the overall pattern of risk remained the same. The largest attenuation was observed in the high adversity group, where the HR went from 4.57 (95% CI [3.98-5.26]) to 3.55 (95% CI [3.06-4.11]). Additional adjustment for SGA did not change the estimates for accident mortality (Supplementary Figure S5).

## Discussion

We examined the association between cumulative exposure to childhood adversities and the risk of accident mortality in early adulthood (16-42 years) using a nationwide Danish cohort of over 1.2 million individuals. We observed 2,366 accidental deaths during follow-up, the majority of which were due to traffic accidents (Supplementary Table S2). We saw a major sex difference in absolute mortality risk with men having higher incidences of all accident types, with men accounting for more than 80% of all cases of accident mortality in this age group. The results demonstrated that children with a history of childhood adversities had higher risk of accidental death in early adulthood across all accident types, with a particularly high risk being observed among the high adversity group. Specifically, for the accident types ‘narcotics and hallucinogens’ and ‘other poisoning’, the high adversity group had a 10 to 13-fold higher risk of mortality compared to the low adversity group.

Overall, the findings of this study are aligned with previous studies finding a higher mortality risk among those who have been exposed to childhood adversity^4,8,9,11,13,16,17,20^. A study looking specifically at accident mortality found a higher risk among those experiencing sibling death in childhood^11^ and a study looking into deaths from poisonings, alcohol, drugs and other substances found a markedly higher risk associated in those with a history of contact with child protection services^21^, though the estimates were lower than in the current study. In contrast to the current study, these studies investigated single adversities in isolation. Assuming that accumulation of adversity puts individuals at higher risk of death, this may explain why these studies found weaker associations. A study assessing the association between trajectory groups of adversity and mortality from substance use found an increased risk across nearly all adversity groups, as we did^9^. However, this study identified differing adversity groups and measured adversities from age 0-7, whereas we measured adversities across the entirety of childhood (years 0-15).

Several studies have established that childhood adversities are associated with hazardous and harmful health behaviors in adolescence and early adulthood, such as alcohol– and substance abuse^1,10,12,22–24^. Toxic stress (i.e. severe, chronic stress from persistent adversity exposure in the absence of supportive adult relations) has been suggested to link childhood adversities with risky health behaviors^12^ and alterations to the neurobiological stress response systems and the brain’s architecture in areas for memory, learning and decision-making^25^. Thus, exposure to childhood adversities may induce toxic stress, which can impact brain architecture and development, causing detrimental changes to individual’s behavioral patterns and stress response. Brain plasticity during the foetal, infant and early childhood makes the brain especially sensitive to the influence of the toxic stress^12^. This suggests a biologically founded hypothesis of varying risk behavior in children and young adults who have experienced different amounts of childhood adversities.

### Strengths and limitations

A principal strength of this study lies in its use of nationwide life-course data from high-resolution registers. This ensures a large, unselected population with clear temporal data on exposures, covariates and outcomes. The high-quality data in the registries has enabled us to reliably and comprehensively measure childhood adversity based on 12 adversities across three dimensions across the entirety of childhood. In addition, the yearly repeated measures of childhood adversities (0-15 years) yield valuable information on both accumulation and timing of adversities. Another particular contribution of this study is that we looked into lethal accidents of various types. By examining distinct types of lethal accident we can identify which subgroups, defined by sex and adversity groups, are at greater risk and when in early adulthood this risk is highest, thus providing more detailed evidence to inform future targeted prevention strategies and policies.

Relying on nationwide objective life-course data on childhood adversity is a major strength of the study, but it also imposes some limitations. For example, only individuals whose parents received formal treatment or medication for alcohol use disorder were classified as exposed to parental alcohol abuse (7%) in this study^5^. National estimates suggest considerable underreporting in clinical records^26^. As a result, the prevalence may be underestimated. In addition, childhood adversities such as sexual abuse and violence are not captured in Danish registers^4^. Exposure to abuse, neglect and maltreatment can be extremely stressful and detrimental for children, and strong associations have been shown between abuse in childhood and later life mortality^27^. However, we expect that due to the clustering of adversities children who experienced these adversities also experienced other adversities and therefore the risk of misclassification due to the absence of this information is reduced.

Another limitation is the risk of misclassification where suicides are registered as accidents. However, in cases where there is insufficient evidence of intent of suicide to distinguish between accidental causes of death and intentional self-harm, coroners or doctors use the ICD-10 codes for ‘undetermined intent’ (Y10-Y34) in cause of death registration^28^, thus limiting the risk of misclassification of suicides as accidental causes of death, making this source of misclassification disregardable. We excluded the few children who emigrated or died before age 16 years, most of which were due to emigration. It is well established that there is increased risk of childhood mortality among those who experienced a high degree of childhood adversity compared with those who did not experience adversities^29^. Therefore, childhood adversities may affect the risk of accident mortality already before the age of 16, which may have caused an underestimation of the true association^30^ between childhood adversities and accident mortality in this study. However, as the mortality rate in the population in this age span is very low^4^, the magnitude of this bias is expected to be minor.

The results from this study emphasize the need for future preventive socio-environmental and socioeconomic efforts to minimize the occurrence of childhood adversities, as well as targeted accidental mortality prevention strategies. For example, traffic accidents among young men would be a major point for intervention, as these constitute the vast majority of all accidental causes of death in young adulthood in Denmark. Furthermore, approaches to interventions focusing on circumstances and factors that ameliorate the detrimental effects of childhood adversities should be strengthened.

## Conclusion

The experience of adversity is common among children but varies in degree. A smaller group of children experience multiple adversity across various dimensions, and these children seem to be particularly vulnerable to the life-course consequences of childhood adversity. We have shown that children who experience childhood adversities, particularly those with high adversity, have a higher risk of dying from accidental causes in early adulthood. This was the case for all adversity groups and in all accident types. Furthermore, we found a considerable sex difference in absolute mortality risk with men having higher incidences of all accident types. Structural and social interventions must be employed to reduce the exposure to childhood adversities, ameliorate the harmful effects of childhood adversities as well as reduce excess risk of mortality due to accidents in early adulthood.

## Supplementary information

The online version contains supplementary material.

## Acknowledgements

We would like to thank J. Bengtsson and A. Rieckmann for their conceptual and statistical assistance during the initial phase of the project.

## Author contributions

Line M. T. Dyhr – Writing, review & editing. Writing – Original draft, Visualization, Methodology, Formal analysis, Conceptualization.

Leonie K. Elsenburg – Writing, review & Editing, Supervision, Project Administration, Methodology, Conceptualization.

Naja H. Rod – Writing, review & editing, Methodology, Conceptualization.

## Data availability

The data material contains sensitive and personally identifiable information, why it cannot be made publicly available. Inquiries about secure access to the DANLIFE data under the stated conditions by the Danish Data Protection Agency can be directed at Naja Hulvej Rod.

## Supplementary material

**Supplementary Table S1.** Childhood adversities, their definition, and the registers informing them Rod et al. (2020) [1]. Further details on the definitions can be found in Bengtsson et al. (2019) [2].

|  | Adversity | Definition | Registers |
| --- | --- | --- | --- |
| Material deprivation | Family poverty | Family income below 50% of the median national family income in a given year | The Income Statistics Register [3] |
|  | Parental long-term unemployment | Parental unemployment for at least 12 months | The Integrated Database for Labour Market Research [4] |
| Loss or threat of loss | Death of a parent | Death of a parent | The Danish Civil Registration System [5] |
|  | Death of a sibling | Death of a sibling | The Danish Civil Registration System [5] |
|  | Parental somatic illness | A parent being diagnosed with one of the diseases included in the Charlson comorbidity index | The Danish National Patient Register [6] |
|  | Sibling somatic illness | Having a sibling being diagnosed with one of the seven somatic illnesses most commonly related to mortality in children aged 0–18 years in Denmark: malignant neoplasm; congenital anomalies of the heart and circulatory system; congenital anomalies of the nervous system; cerebral palsy; epilepsy; cardiomyopathy; congenital disorders of lipid metabolism | The Danish National Patient Register [6] |
| Family dynamics | Foster care | Placement in out-of-home care | The Register of Support for Children and Adolescents [7] |
|  | Parental psychiatric illness | Parental admission for at least 1 day to a psychiatric hospital or ward with a primary diagnosis related to psychiatric illness (excluding primary diagnoses related to alcohol and drug abuse) | The Danish Psychiatric Central Research Register [8]; The Danish National Patient Register [6] |
|  | Sibling psychiatric illness | Having a sibling admitted for at least 1 day to a psychiatric hospital or ward with a primary diagnosis related to psychiatric illness | The Danish Psychiatric Central Research Register [8]; The Danish National Patient Register [6] |
|  | Parental alcohol abuse | Parental diagnosis with a disease related to alcohol abuse or buying a prescribed drug used in treatment of alcohol dependence | The Danish Psychiatric Central Research Register [8]; The Danish National Patient Register [6]; The Danish National Prescription Registry [9] |
|  | Parental drug abuse | Parental diagnosis with a disease related to drug abuse or buying a prescribed drug used in treatment of drug dependence | The Danish Psychiatric Central Research Register [8]; The Danish National Patient Register [6]; The Danish National Prescription Registry [9] |
|  | Maternal separation | The mother no longer sharing address with a partner | The Danish Civil Registration System [5] |

**Note:** This table has been reproduced from supplementary materials of the study by: Rod NH, Bengtsson J, Elsenburg LK, Taylor-Robinson D, Rieckmann A. *Hospitalisation patterns among children exposed to childhood adversity: a population-based cohort study of half a million children*. *Lancet Public Health*, 2021; published online September 29. http://dx.doi.org/10.1016/S2468-2667(21)00158-4

---

**Supplementary Figure S1.**
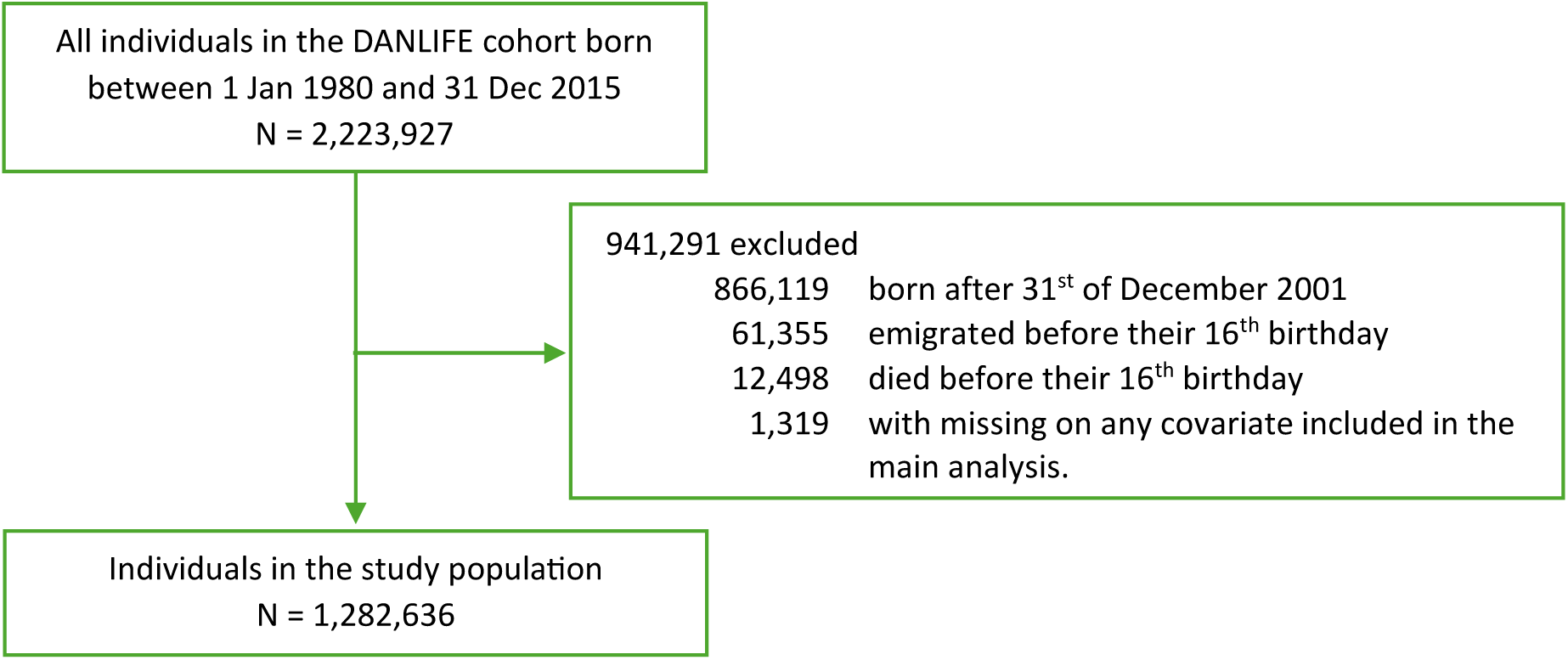
Flowchart of the study population. The study population was followed up for a total of 18,483,241 person-years. As the Danish emigration register is updated back in time, 3,326 individuals in our study population turned out to have emigrated before their 16^th^ birthday and thus had no follow-up time.

**Supplementary Table S2.** Frequency of accident mortality according to accident type and sex.

| Accident type | Men, N (%*) | Women, N (%*) | Total, N (%**) |
| --- | --- | --- | --- |
| Traffic accidents | 1057 (80.0%) | 265 (20.0%) | 1,322 (55.9%) |
| Narcotics & hallucinogenics | 299 (80.0%) | 75 (20.0%) | 374 (15.8%) |
| Other poisoning accidents | 309 (83.0%) | 63 (17.0%) | 372 (15.7%) |
| Other accident types | 252 (84.6%) | 46 (15.4%) | 298 (12.6%) |
| Total | 1,917 (81.0%) | 449 (19.0%) | 2,366 |

\*Row percentages. Shows the proportion within the total cases of the specific accident type
\*\* Column percentages. Shows the proportion within total cases of all accident types

**Supplementary Figure S2.**
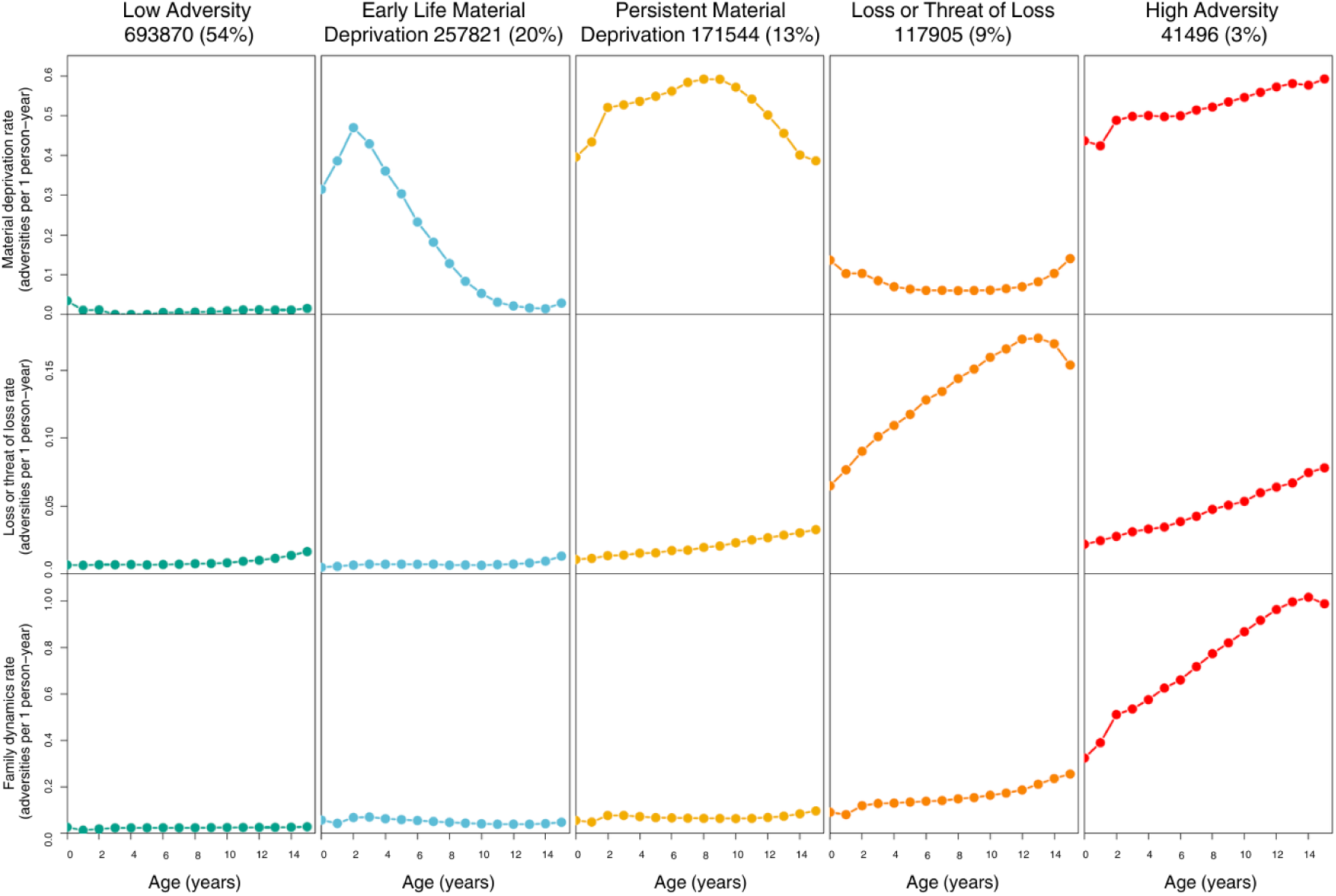
Annual rates of 12 childhood adversities between age 0-15 years, across the three dimensions: material deprivation, loss or threat of loss, and family dynamics. Rates are given as number of events per 1 person-year. The five columns represent the childhood adversity trajectory groups: low adversity (green), early life material deprivation (blue), persistent material deprivation (yellow), loss or threat of loss (orange), and high adversity (red), as identified by Rod et al. (2020, 2021). Percentages indicate proportion of children in the DANLIFE population belonging to each of the five trajectory groups.

**Supplementary Figure S3.**
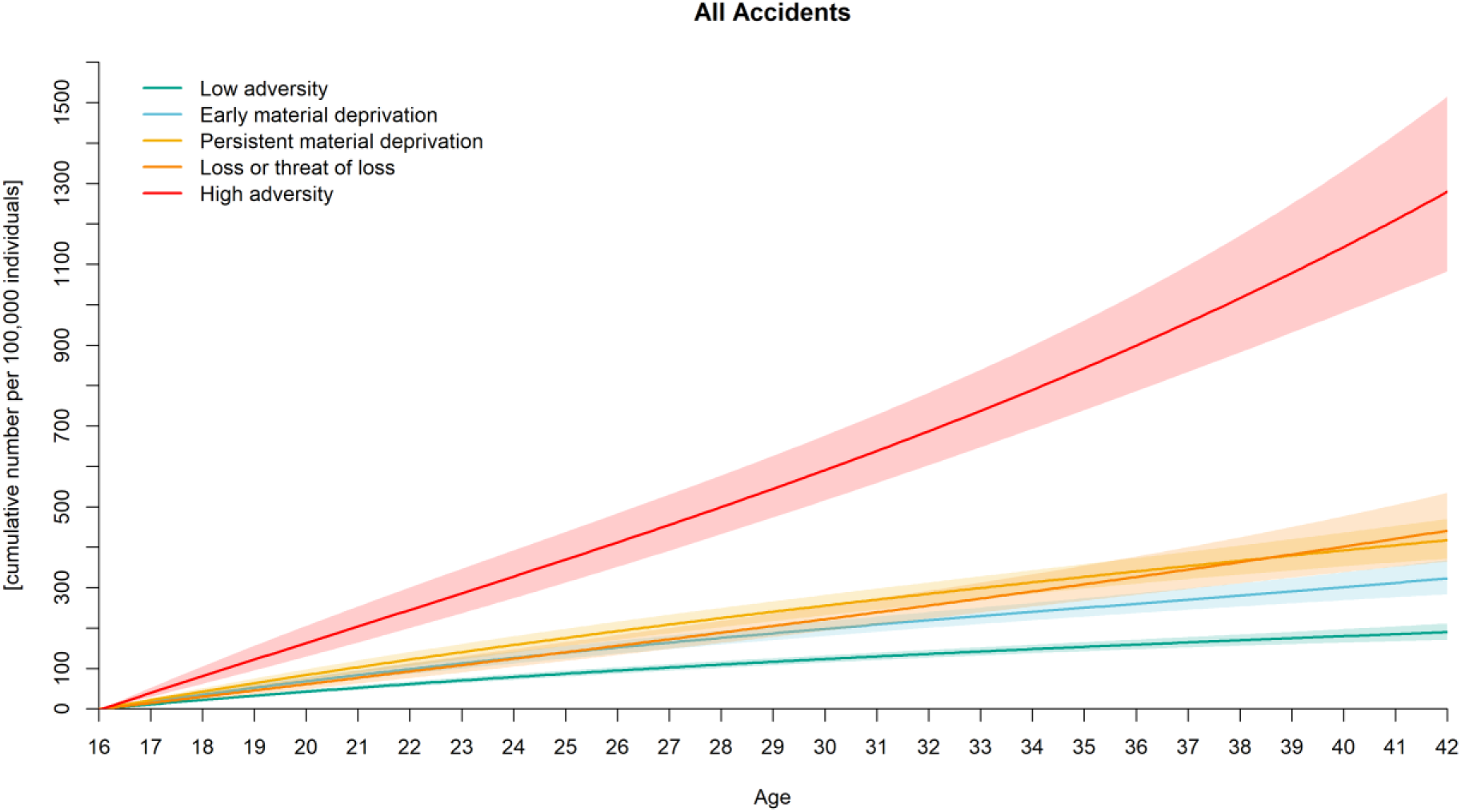
Cumulative risk of all lethal accidents per 100,000 individuals in each of the five adversity trajectory groups.

**Supplementary Figure S4.**
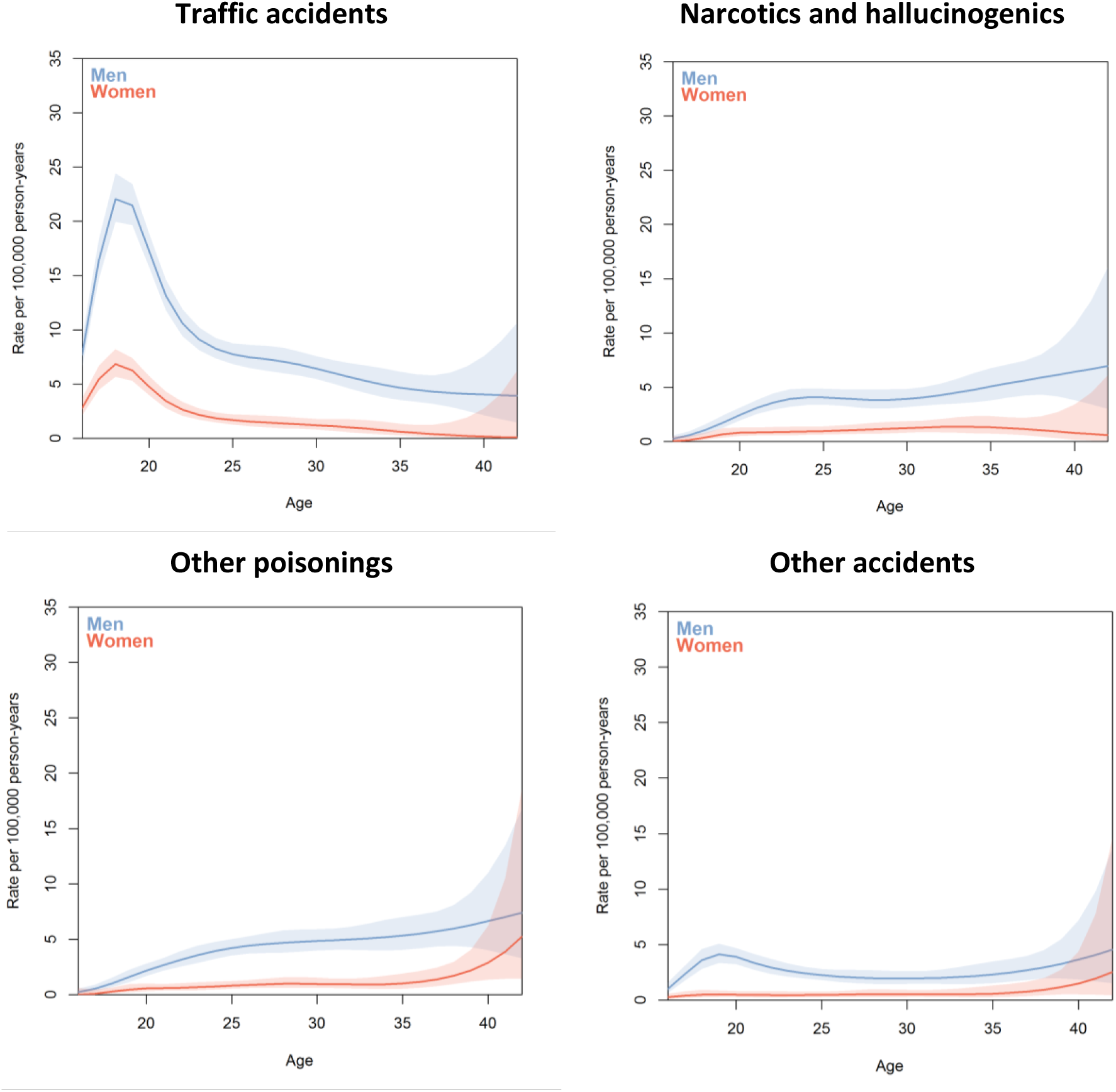
Age-specific incidence rate of accident type specific mortality per 100,000 person-years according to sex. Poisson models were fitted with splines (df = 6).

**Supplementary Table S3.**
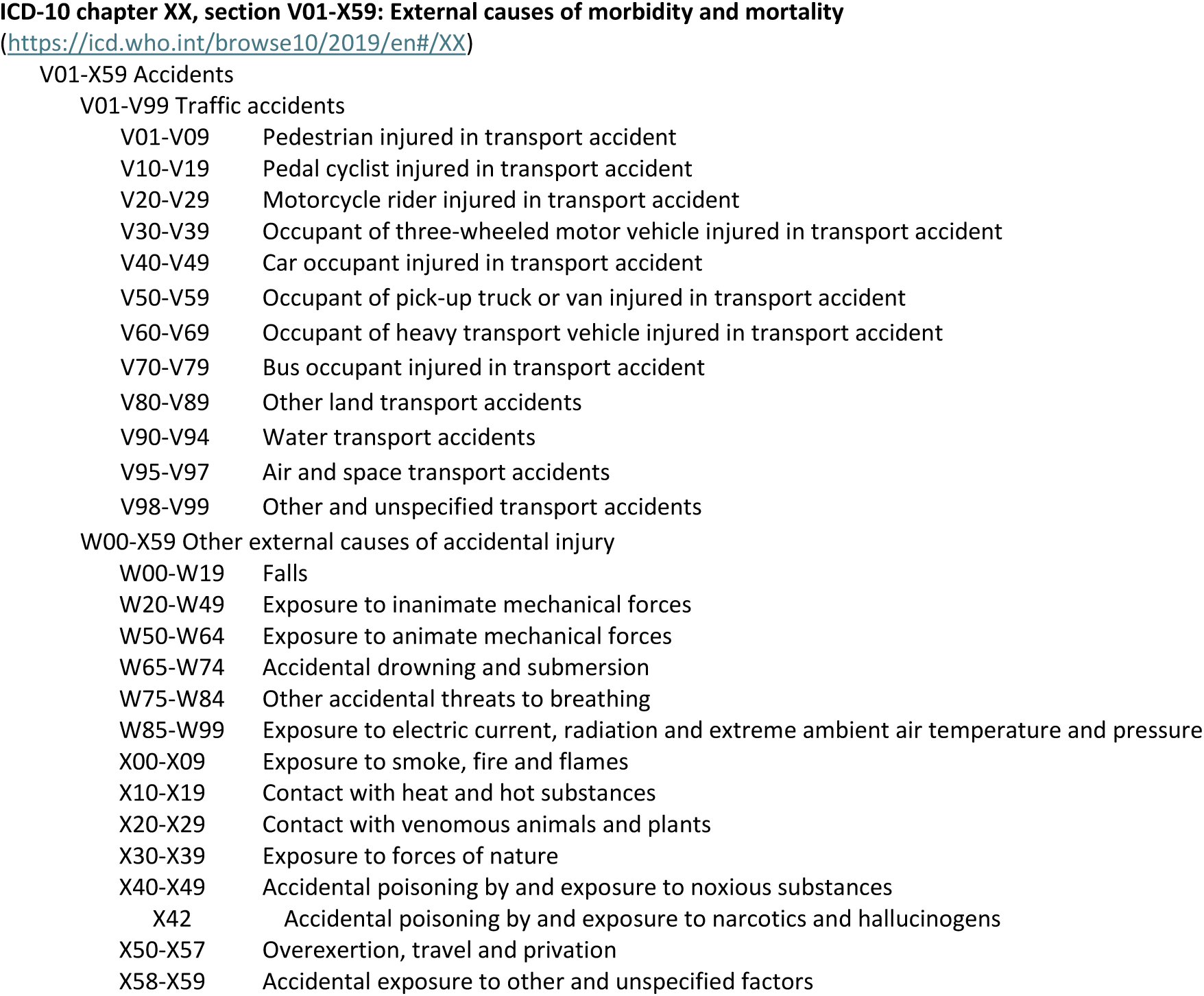
Overview of the ICD-10 classification of chapter XX, subsection of external causes of morbidity and mortality.

V01-V99 Traffic accidents
|  |  |
| --- | --- |
| V01-V09 | Pedestrian injured in transport accident |
| V10-V19 | Pedal cyclist injured in transport accident |
| V20-V29 | Motorcycle rider injured in transport accident |
| V30-V39 | Occupant of three-wheeled motor vehicle injured in transport accident |
| V40-V49 | Car occupant injured in transport accident |
| V50-V59 | Occupant of pick-up truck or van injured in transport accident |
| V60-V69 | Occupant of heavy transport vehicle injured in transport accident |
| V70-V79 | Bus occupant injured in transport accident |
| V80-V89 | Other land transport accidents |
| V90-V94 | Water transport accidents |
| V95-V97 | Air and space transport accidents |
| V98-V99 | Other and unspecified transport accidents |
| W00-X59 Other external causes of accidental injury |  |
| W00-W19 | Falls |
| W20-W49 | Exposure to inanimate mechanical forces |
| W50-W64 | Exposure to animate mechanical forces |
| W65-W74 | Accidental drowning and submersion |
| W75-W84 | Other accidental threats to breathing |
| W85-W99 | Exposure to electric current, radiation and extreme ambient air temperature and pressure |
| X00-X09 | Exposure to smoke, fire and flames |
| X10-X19 | Contact with heat and hot substances |
| X20-X29 | Contact with venomous animals and plants |
| X30-X39 | Exposure to forces of nature |
| X40-X49 | Accidental poisoning by and exposure to noxious substances |
| X42 | Accidental poisoning by and exposure to narcotics and hallucinogens |
| X50-X57 | Overexertion, travel and privation |
| X58-X59 | Accidental exposure to other and unspecified factors |

**Supplementary Figure S5.**
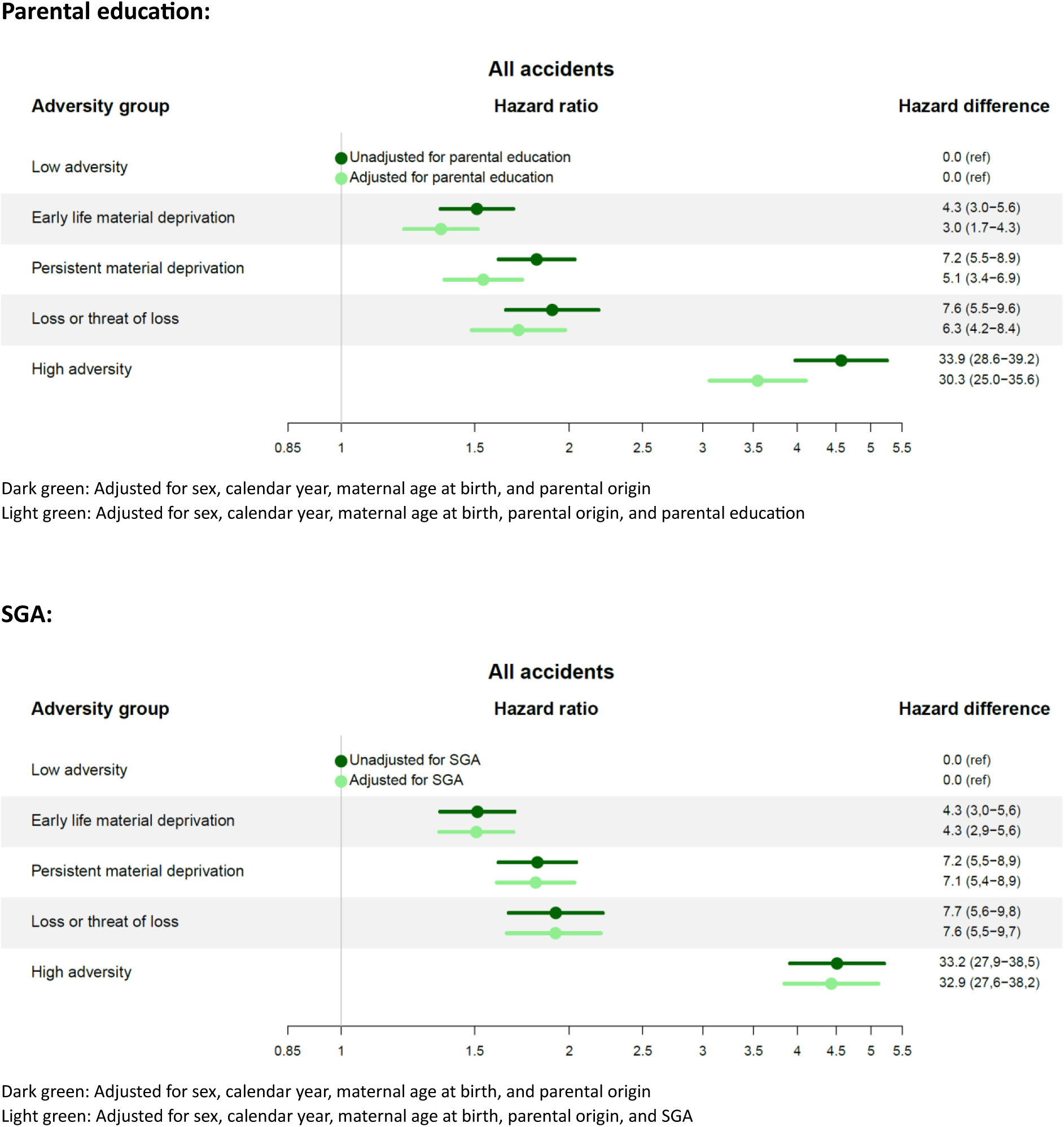
Sensitivity analyses of parental education and small for gestational age (SGA), respectively. Hazard ratios and hazard differences per 100,00 person-years for accident mortality. Analyses were restricted to individuals with full information on parental education. Both models were adjusted for sex, calendar year, maternal age at birth, and parental origin, with one model additionally adjusting for parental education and the other model additionally adjusting for SGA.

## References

1. Hughes, K. et al. The effect of multiple adverse childhood experiences on health: a systematic review and meta-analysis. Lancet Public Health 2, e356–366 (2017).

2. Jackisch, J., Ploubidis, G. B. & Gondek, D. Does time heal all wounds? Life course associations between child welfare involvement and mortality in prospective cohorts from Sweden and Britain. SSM – Popul. Health 14, 100772 (2021).

3. Kelly-Irving, M. et al. Adverse childhood experiences and premature all-cause mortality. Eur. J. Epidemiol. 28, 721–34 (2013).

4. Rod, N. et al. Trajectories of childhood adversity and mortality in early adulthood: a population-based cohort study. The Lancet 396, 489–497 (2020).

5. Bengtsson, J., Dich, N., Rieckmann, A. & Rod, N. H. Cohort profile: the DANish LIFE course (DANLIFE) cohort, a prospective register-based cohort of all children born in Denmark since 1980. BMJ Open 9, e027217 (2019).

6. Felitti, V. J. et al. Relationship of Childhood Abuse and Household Dysfunction to Many of the Leading Causes of Death in Adults The Adverse Childhood Experiences (ACE) Study. Am. J. Prev. Med. 14, 245–258 (1998).

7. Kuh, D., Ben-Shlomo, Y., Lynch, J., Hallqvist, J. & Power, C. Life course epidemiology. J. Epidemiol. Community Health 57, 778–783 (2003).

8. Bhattarai, A., Dimitropoulos, G., Bulloch, A. G. M., Tough, S. C. & Patten, S. B. Association between childhood adversities and premature and potentially avoidable mortality in adulthood: a population-based study. BMC Public Health 23, 2036 (2023).

9. Govender, T. et al. Adverse childhood experiences and risk of suicide and substance-related mortality through middle adulthood. J. Affect. Disord. 369, 1201–1208 (2025).

10. Grummitt, L. R. et al. Association of Childhood Adversity With Morbidity and Mortality in US Adults: A Systematic Review. JAMA Pediatr. 175, 1269–1278 (2021).

11. Li, J. et al. Mortality after parental death in childhood: a nationwide cohort study from three Nordic countries. PLoS Med. 11, e1001679 (2014).

12. Shonkoff, J. P. & Garner, A. S. The Lifelong Effects of Early Childhood Adversity and Toxic Stress. Pediatrics 129, e232–e246 (2012).

13. Yu, J. et al. Adverse childhood experiences and premature mortality through mid-adulthood: A five-decade prospective study. Lancet Reg. Health Am. 15, 100349 (2022).

14. Yu, Y. et al. Association of Mortality With the Death of a Sibling in Childhood. JAMA Pediatr. 171, 538–545 (2017).

15. Zheng, L. et al. Association between adverse childhood experiences and mortality: A systematic review and meta-analysis. Psychiatry Res. 343, 116275 (2025).

16. Martikainen, P., et al. The Changing Contribution of Childhood Social Characteristics to Mortality: A Comparison of Finnish Cohorts Born in 1936-50 and 1961-75. vol. 49 (2020).

17. Rostila, M., Berg, L., Saarela, J., Kawachi, I. & Hjern, A. Experience of Sibling Death in Childhood and Risk of Death in Adulthood A National Cohort Study From Sweden. Am. J. Epidemiol. 185, 1247–1254 (2017).

18. Schmidt, M., Pedersen, L. & Sørensen, H. T. The Danish Civil Registration System as a tool in epidemiology. Eur. J. Epidemiol. 29, 541–549 (2014).

19. Helweg-Larsen, K. The Danish Register of Causes of Death. Scand. J. Public Health 39, 26–29 (2011).

20. Yu, Y. et al. Association of Mortality With the Death of a Sibling in Childhood. JAMA Pediatr. 171, 538–545 (2017).

21. Segal, L. et al. Child Maltreatment and Mortality in Young Adults. Pediatrics 147, e2020023416 (2021).

22. Espeleta, H. C., Brett, E. I., Ridings, L. E., Leavens, E. L. S. & Mullins, L. L. Childhood adversity and adult health-risk behaviors: Examining the roles of emotion dysregulation and urgency. Child Abuse Negl. 82, 92–101 (2018).

23. Giordano, G. N., Ohlsson, H., Kendler, K. S., Sundquist, K. & Sundquist, J. Unexpected adverse childhood experiences and subsequent drug use disorder: A Swedish population study (1995–2011). Addiction 109, 1119–1127 (2014).

24. Jones, M. S., Pierce, H. & Shafer, K. Gender differences in early adverse childhood experiences and youth psychological distress. *Forthcom*. J. Crim. Justice 1–11 (2022).

25. Kashef, Z. Exposure to Toxic Stress in Childhood Linked to Risky Behavior and Adult Disease. Yale Nurs. Matters Fall, 6–9 (2015).

26. Sundhedsstyrelsen. Forebyggelsespakke – mental sundhed. (2018).

27. Chen, E., Turiano, N. A., Mroczek, D. K. & Miller, G. E. Associations of reports of childhood abuse and all-cause mortality rates in women. JAMA 73, 920–927 (2016).

28. WHO. ICD-10 Version 2019. Event of undetermined intent (Y10-Y34). (2022).

29. Grey, H. R., Ford, K., Bellis, M. A., Lowey, H. & Wood, S. Associations between childhood deaths and adverse childhood experiences. Child Abuse Negl. 90, 22–31 (2019).

30. Hernán, M. A., Hernández-Dias, S. & Robins, J. M. A structural approach to selection bias. Epidemiol Camb Mass 615–625 (2004).

## References

1. Rod, N.H., et al., Supplement to: Rod NH, Bengtsson J, Elsenburg LK, Taylor-Robinson D, Rieckmann A. Hospitalisation patterns among children exposed to childhood adversity: a population-based cohort study of half a million children. Lancet Public Health, 2021.

2. Bengtsson, J., A. Rieckmann, and N.H. Rod, Cohort profile: the DANish LIFE course (DANLIFE) cohort, a prospective register-based cohort of all children born in Denmark since 1980. BMJ Open, 2019. 9: p. e027217.

3. Baadsgaard, M. and J. Quitzau, Danish registers on personal income and transfer payments. Scandinavian Journal of Public Health, 2011. 39(7 Suppl): p. 103–105.

4. Petersson, F., M. Baadsgaard, and L. Thygesen, Danish registers on personal labour market affiliation. Scandinavian Journal of Public Health, 2011. 39(7 Suppl): p. 95–98.

5. Schmidt, M., L. Pedersen, and H.T. Sørensen, The Danish Civil Registration System as a tool in epidemiology. European Journal of Epidemiology, 2014. 29: p. 541–549.

6. Schmidt, M., et al., The Danish National Patient Registry: A review of content, data quality, and research potential. Clinical Epidemiology, 2015. 7: p. 449–490.

7. DST. Description of the Register of Support for Children and Adolescents [in Danish]. 2021 [cited 2021 June 7]; Available from: https://www.dst.dk/da/TilSalg/Forskningsservice/Dokumentation/hoejkvalitetsvariable/stoette-til-udsatteboern-og-unge.

8. Mors, O., G. Perto, and P. Mortensen, The Danish Psychiatric Central Research Register. Scandinavian Journal of Public Health, 2011. 39(7 Suppl): p. 54–57.

9. Pottegård, A., et al., Data resource profile: The Danish National Prescription Registry. International Journal of Clinical Epidemiology, 2017. 46(3): p. 798.

